# DeepEXPOKE: A Deep Learning Framework with Polygenic Risk Scores as Knockoffs for Deconvoluting Genetic and Non-Genetic Exposure Risks in Sepsis and Coronary Heart Disease

**DOI:** 10.1101/2024.10.15.24315572

**Authors:** Aditya Sriram, Rebecca Bohn, Kate Kernan, Joseph Carcillo, Soyeon Kim, Hyun Jung Park

## Abstract

The exposome refers to the totality of environmental, behavioral, and lifestyle exposures an individual experiences throughout one’s lifetime. Due to the modifiability of exposures, identifying the risk exposures on a disease is crucial for effective intervention and prevention of the disease. However, traditional analytical methods struggle to capture the complexities of exposome data: nonlinear effects, correlated exposures, and potential interplay with genetic effects. To address these challenges and accurately estimate exposure effects on complex diseases, we developed DeepEXPOKE, a deep learning framework integrating two types of knockoff features: statistical knockoffs (statKO) and polygenic risk score as knockoffs (PRSKO). DeepEXPOKE-statKO controls exposure correlation and DeepEXPOKE-PRSKO isolates genetic effects, while both can capture nonlinear effects. We applied DeepEXPOKE to predict outcomes of two significant diseases with distinct etiology and clinical presentation: sepsis and coronary heart disease (CHD), demonstrating its performance in comparison to existing machine learning methods. Furthermore, both DeepEXPOKE-PRSKO and DeepEXPOKE-statKO identified metabolites such as glucose and triglycerides as risk factors for sepsis and suggested that their effects are primarily at the non-genetic level, consistent with the role of metabolites in responding to environmental factors. Additionally, DeepEXPOKE-PRSKO uniquely identified asthma as a sepsis risk factor and suggested its effect is partially at the genetic level, offering insights into the conflicting associations observed between the genome data studies and patient data analysis regarding asthma and sepsis risk. Overall, DeepEXPOKE offers a novel DNN approach for identifying and interpreting exposure risk factors, advancing our understanding of complex diseases.

## INTRODUCTION

The exposome refers to the totality of environmental, behavioral, and lifestyle exposures an individual experiences throughout their lifetime^1^. Thus, they are modifiable primarily through changes in those aspects of one’s life. Due to the modifiability, identifying the risk exposures is crucial to effectively intervene and prevent complex diseases^2–4^. Recently, population-based health databases have been organized to provide a wide range of exposures collected from many subjects, together with their genome data. For example, the UK Biobank (UKBB) provides a diverse range of exposures of approximately 500,000 participants from the United Kingdom^5,6^. Thus, an emerging task is to effectively explore the large databases and identify risk exposures linked to complex diseases. Especially, to estimate the modifiable portion of the risk separated from genetic influences, several analytical approaches considering both genetic variants and exposures can be applied^7^. First, regression models are used to assess interaction terms pairing genes and environmental factors^8–10^. For example, Liu et al. explored how the interactions between particular genetic variants and environmental exposures shapes brain structure and function^8^. Second, polygenic risk score (PRS), combined with environmental data, improves disease risk prediction and examines the interactions between genetic and environmental factors. For example, Khera et al. demonstrated that integrating PRS with lifestyle factors, such as diet and exercise, better stratify cardiovascular disease patients for Cox proportional-hazard models ^11^. Third, Mendelian Randomization (MR) leverages genetic variants as instrumental variables to infer causal relationships between exposures and outcomes. This approach has been particularly useful in distinguishing causality from correlation in observational studies^7, 12, 13^.

While the studies highlight the significant contributions of the interactions between genetic factors and exposures, multiple limitations have been discussed of the current studies for accurate estimation of exposure risk^14, 15^ (**Fig. 1A**). First, exposures affect outcomes in highly nonlinear fashions due to complex interaction patterns that involve the threshold effect, inverse relationship, or saturation effect^16^.

**Figure 1.**
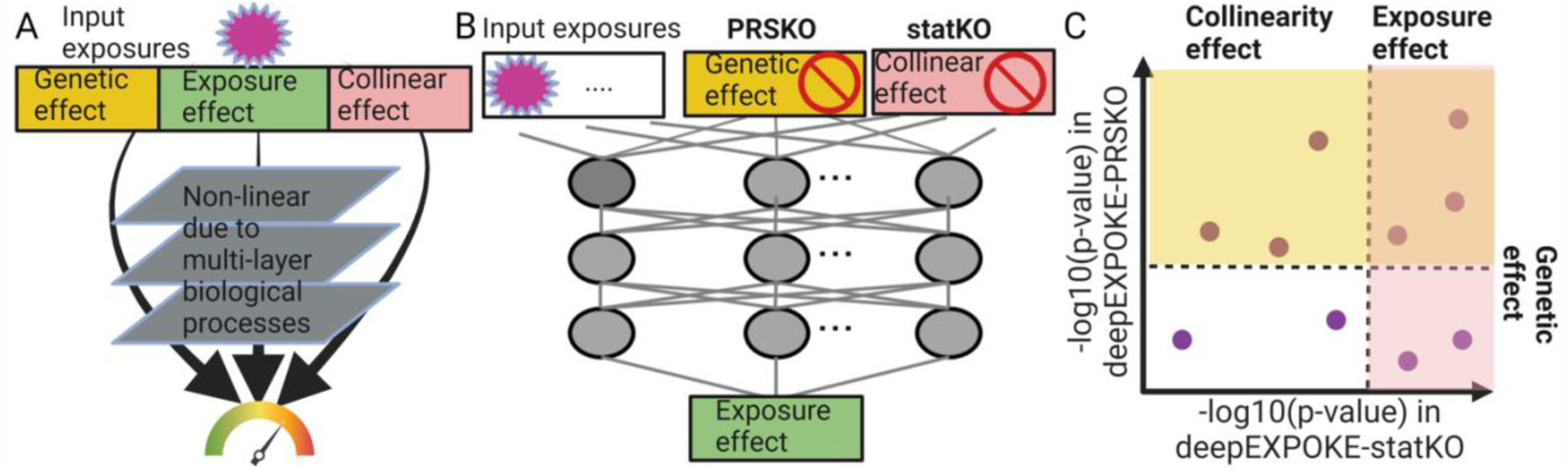
DeepEXPOKE identifies exposure risk factors in the DNN framework with novel PRS knockoff features. **A)** Illustration showing how an exposure exerts effect by passing the genetic, exposure, and collinear effect through multi-layer biological processes. **B)** deepEXPOKE architecture that estimates the exposure effect by controlling the genetic and/or collinear effect via PRS knockoffs (KO) and statistical (stat) knockoffs, respectively. **C)** Illustration showing the effect size in PRS and/or stat KO indicates collinearity, genetic, or exposure effect. Note that if an exposure effect is identified significant by both PRSKO and statKO, then the effect is estimated to be at the exposure level.

However, previous studies have been based on linear models, such as regression^17–20^. Second, exposures are often correlated with each other^5^, making it difficult to attribute observed health outcomes to specific exposures. For example, if both poor diet and lack of exercise are both linked to a disease, current methods are not designed to separate their effects, making it difficult to determine whether to focus on diet or exercise^6^. Third, genetic effects often influence a person’s likelihood of engaging in certain exposures, such as dietary choices or physical activity^21^. If these exposures impact health, it is challenging for existing methods to separate their direct effects from genetic influences. MR methods help separate effects, but mostly based only on linear models, and thus not able to capture the nonlinear effects. The only deep neural network (DNN) model in MR approach, DeepMR^22^, is to link mutagenesis marks in a small genomic region to a related trait. It is not suitable for genome-exposome data analysis, so it was not considered in this manuscript.

To address each of the limitations, we propose a novel <u>deep</u> learning for <u>expo</u>some analysis using <u>k</u>nockoff <u>e</u>stimation (deepEXPOKE) (**Fig. 1B**). First, to model the nonlinear interaction patterns between disease outcomes and their exposures, we will employ multiple layers of nonlinear activators in the deep neural network (DNN) framework. We previously showed that this DNN approach successfully validates known nonlinear interactions and identifies novel therapeutic targets for complex diseases, such as sepsis and breast cancer^23^. Second, to control correlations between exposures, we propose to employ knockoff features in the DNN approach. Knockoff features are synthetic and noisy copies of the input variables, which 1) resemble the correlation structure of the input variables but 2) are conditionally independent of the outcome, given the input variables^24^. Due to these two mathematical conditions that control the correlation structure of the input variables, knockoff features have been effective in dealing with variable correlation issues in DNN approaches^23, 25^, although they have not been applied to exposure data yet. Third, to separate exposure effects from genetic influence, we propose a novel type of knockoff based on polygenic risk scores (PRS). Standard knockoffs are statistically generated and thus do not represent the genetic influence on the exposures. On the other hand, PRS, the aggregate effect of multiple genetic variants to a specific trait, represents a baseline level of genetic influence while satisfying the two mathematical conditions to be knockoff given above. First, if two exposures share genetic factors, they will correlate in PRS, satisfying the first mathematical condition to be knockoff.

Second, PRS or a subset of genetic variants in the PRS model are used to infer causal relationships between exposures and health outcomes^26^. This is because PRS is independent of the outcomes conditioned on the exposures, which demonstrates that PRS satisfies the second mathematical condition to be a knockoff variable.

Based on these ideas, we developed deepEXPOKE with two types of knockoffs: statistical knockoff (statKO) and PRS knockoff (PRSKO). We tested deepEXPOKE on two complex diseases with distinct clinical presentation, sepsis and coronary heart disease (CHD) (**S. Fig. 1**). Sepsis, accounting for 20% of all deaths worldwide^27^, is initiated by viral/bacterial infection and involves dysfunctional immune system.

Exposures such as nutritional deficiencies or stress can raise sepsis risk^28, 29^, while genetic factors can also influence exposures. On the other hand, coronary heart disease (CHD), which involves blockage of the coronary arteries, is influenced by exposures, such as diet, smoking, and physical activity^30^. Using the disease data downloaded from UKBB, we will validate PRS as a type of knockoff based on the mathematical properties. Then, we will evaluate DeepEXPOKE in predicting sepsis and CHD incidence and mortality in comparison to other machine learning methods. In the process, we will also identify individual exposure risk factors and interpret their roles in clinical outcomes. DeepEXPOKE also offers alternative explanations for previous inconsistencies, potentially arising from biases such as exposure correlation or genetic effects. This includes complex relationships like the ’hypertension paradox^31^, where hypertension is paradoxically associated with better outcomes in certain contexts. Together, deepEXPOKE provides the first deep-learning framework to elucidate the exposure risk to complex diseases by addressing collinearity and genetic factors.

## RESULTS

### DeepEXPOKE identifies exposure risk factors in the DNN framework with novel PRS knockoff features

To control for correlation with other exposures and underlying genetic effects when estimating exposure risk for complex diseases, deepEXPOKE incorporates a DNN framework with two different types of knockoff features: statistical knockoffs (statKO) and PRS as knockoffs (PRSKO) (**Fig. 1B**). The statistical knockoffs are constructed to mimic the correlation structure between input variables^24^, while PRS values are used to estimate genetic contributions to the outcome. Since both correlation and genetic effects can influence disease risk through complex, nonlinear pathways across multiple biological layers—such as the genomic, transcriptomic, and proteomic levels—DeepEXPOKE integrates these knockoff features into a DNN model. DNNs utilize multiple layers of nonlinear activators to capture nonlinear patterns from data^23^. Specifically, DeepEXPOKE generates both types of statistical and PRS knockoff for each input feature as a novel type of knockoff and pairs them with the input feature to feed into the DNN. After training on these input-knockoff pairs to predict outcomes, DeepEXPOKE estimates the nonlinear effect sizes using the trained knockoff values, as the trained knockoff values capture the contribution of collinearity or underlying genetic effects to the prediction, respectively (**Fig. 1C**). Specifically, risk factors identified both by deepEXPOKE-PRSKO and deepEXPOKE-statKO exert effect not through underlying genetic influences and collinearity, and thus are exposure-level risk factors. In the same sense, those only by deepEXPOKE-PRSKO exert effect not through underlying genetic influences, affecting by collinearity and/or at the exposure level. However, if they are at the exposure level, then deepEXPOKE-PRSKO should also identify their risk, suggesting they likely exert their effect by collinearity (correlation with other exposures). Lastly, in the same sense, those only by deepEXPOKE-statKO exert effect by genetic influence.

While statistical knockoffs have been applied in DNN frameworks for variable selection and causal inference^23, 25^, we are the first to propose their use in addressing exposure collinearity to our knowledge.

More importantly, the implementation of PRS as a knockoff within the DNN framework is a novel approach that, to our knowledge, has not been explored in any prior research. Overall, DeepEXPOKE is a scientifically innovative and clinically valuable deep learning framework designed to distinguish how exposure effects contribute to phenotypes. This model not only deepens our understanding of complex traits but also holds significant clinical potential by identifying more precise biomarkers and therapeutic targets to address disease management and treatment.

### PRS satisfies the mathematical properties to be a knockoff variable

To validate PRS as a novel type of knockoff, we selected 3,103 participants with history of sepsis, based on established ICD-10 codes (see Methods), and 79,791 healthy controls from UKBB. We then identified 42 clinical and epidemiological input exposures reflecting general health status unaffected by temporal factors (see Methods, **S. Fig. 1**). For each exposure, we estimated a PRS for each input exposure using the pgsc_calc^32^ (see Methods) method and generated statistical knockoffs using the ‘model-X’ framework across all UKBB samples^24^. To assess whether the first mathematical condition in the PRS values was satisfied—correlation with the input values for representing an outcome—we evaluated the correlation between the PRS values and the outcome and compared it with the correlation between the input exposure values and the outcome (**Fig. 2A**). The positive relationship (0.023) observed supports the expectation that knockoffs should reflect the correlation between input exposures and the outcome. To compare it with an established statistical knockoff, we performed the same analysis with the model-X statistical knockoffs to find that the PRS values show a more conducive correlation structure between knockoff and outcome for sepsis incidence than the statistical knockoff values (-0.019). To further validate PRS, we performed the same comparison analysis on the sepsis mortality with 163 non-survivors from 3,181 survivors with sepsis. While ‘model-X’ knockoffs did show a positive correlation with the outcome this time (0.18), the PRS knockoffs showed a stronger positive trend (0.27), demonstrating that PRS satisfies the first mathematical condition as a knockoff, often in an improved manner than the statistical knockoff. To ensure its generalizability beyond sepsis, we performed the same analysis on CHD incidence with 75,184 healthy controls and 7,710 UKBB subjects with CHD history and on CHD mortality with 122 non-survivors from 7,588 CHD cases (**S. Fig. 2A, B**). The PRS knockoffs demonstrate a strong positive correlation trend for both CHD incidence and mortality (0.002 and 0.098, respectively), similar to the statistical knockoff values (0.015 and 0.014, respectively), indicating comparable performance in these contexts.

**Figure 2.**
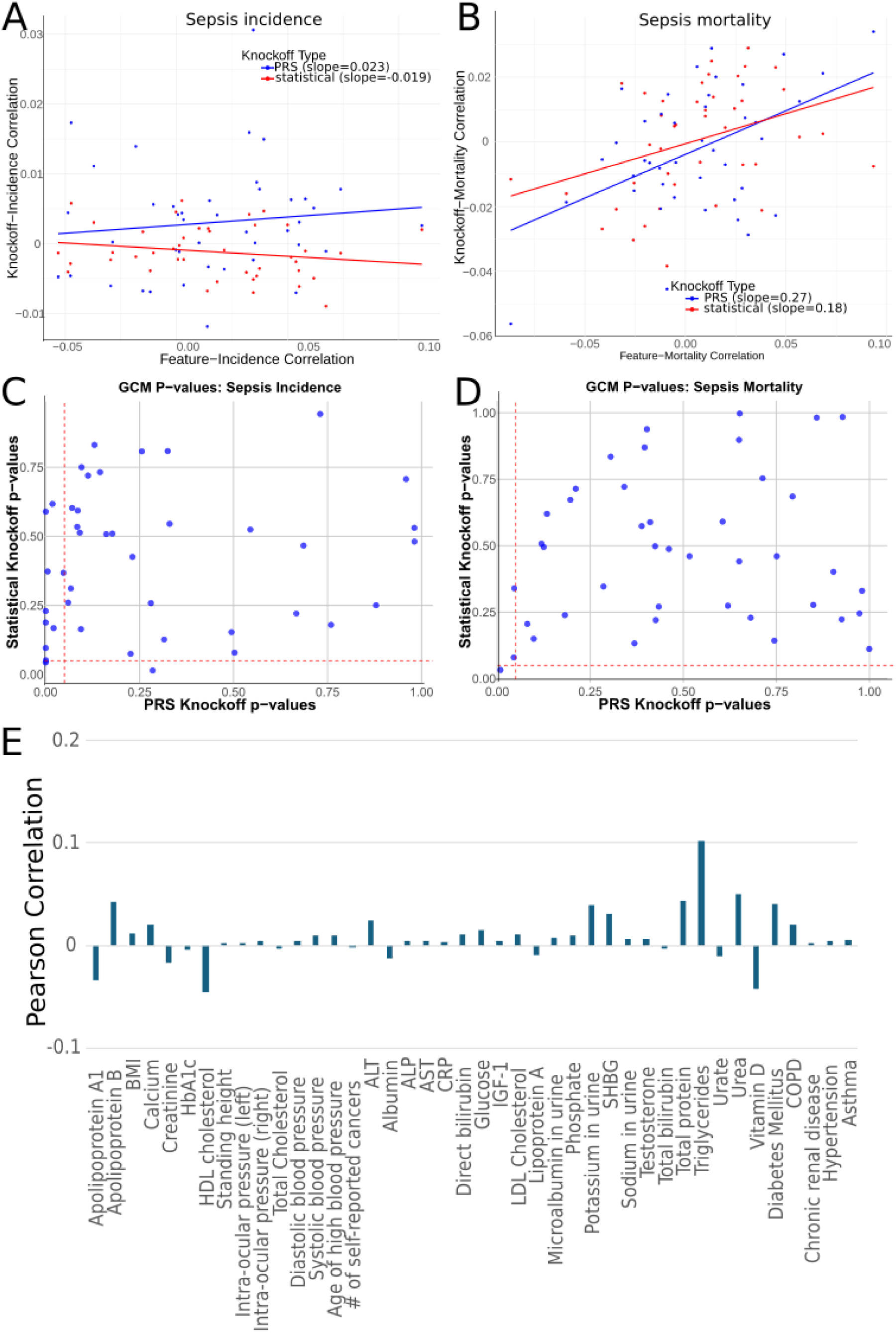
PRS satisfies the mathematical properties to be knockoff. **A)** Scatterplot of correlation values for knockoff variables and sepsis incidence plotted against the correlation values for input exposures and sepsis incidence. **B)** Scatterplot of correlation values for knockoff variables and sepsis mortality plotted against the correlation values for input exposures and sepsis mortality. In A) and B), the blue points and regression line represent the data for PRS knockoffs, and the red points and regression line represent the data for statistical knockoffs. **C)** P-values for the GCM test for conditional independence for the sepsis incidence phenotype for PRS knockoffs (x-axis) and statistical knockoffs (y-axis). **D)** P-values for the GCM test for conditional independence for the sepsis mortality phenotype for PRS knockoffs (x-axis) and statistical knockoffs (y-axis). In C) and D), red dashed lines represent significance levels of 0.05. Points above the horizontal red dashed line exhibit conditional independence in the construct of PRS knockoffs, and points to the right of the vertical red dashed line exhibit conditional independence in the construct of statistical knockoffs. **E)** Pearson correlation coefficient values were calculated between the statistical and PRS knockoff variables for each of our 42 input exposures.

The second condition posits that knockoff features should be independent of the outcome when conditioned on their corresponding input features. To assess this for PRS, we ran generalized covariance measure (GCM), a statistical test for conditional independence that evaluates the normalized empirical covariance between residuals from regression models. Running GCM on the PRS knockoffs with an input exposure and sepsis incidence outcome (**Fig. 2C**), we find that most exposures (80%, 34 out of 42) did not reject the null hypothesis of conditional independence (p-value > 0.05), indicating conditional independence and satisfying the second condition. In the same experiment with sepsis mortality outcome (**Fig. 2D**), even more exposures (95.2%, 40 out of 42) demonstrated the expected conditional independence, comparable to those obtained with statistical knockoffs (95.2% and 97.6%, respectively, **S. Table 2**). Further, the PRS knockoffs exhibit a consistent performance of conditional independence in CHD (**S. Fig. 2C, 2D**), supporting their generalizability.

While PRS serves as a legitimate knockoff as statistical KO for complex diseases, PRS is also expected to capture distinct aspects of the exposure relationships with genetic predisposition, differing from what is represented by statKO. To explore this distinction, we calculated Pearson’s correlation coefficient between the PRSKO and statKO across all the UKBB subjects we had available for each exposure (**Fig. 2E**). The generally low correlation (all below 0.12) indicates that PRSKO and statKO provide distinct and non-redundant information regarding exposure effects. Altogether, these results show that PRS is a valid type of knockoff with inherent biological context across sepsis and CHD, complex diseases with varying etiologies.

### DeepEXPOKE accurately predicts outcomes by selecting important exposures

To comprehensively evaluate the performance, we developed both DeepEXPOKE-statKO and DeepEXPOKE-PRSKO models using the 42 input exposures and applied each respective method to the UKBB sepsis incidence data with 5-fold cross validation. We compared these models to other established machine learning methods of different algorithms, XGBoost and Random Forest to compare sepsis case prediction performance. XGBoost (Extreme Gradient Boosting) is an ensemble learning method based on decision trees^33^. It builds multiple decision trees sequentially, where each tree corrects the errors of the previous one. While Random Forest is also an ensemble method^34^, it consists of multiple decision trees constructed independently from different random subsets of the data. The final prediction is made by majority voting. We avoided developing machine learning models that are not straightforward to perform variable selection with, such as support vector classifier, as our interest is to further identify exposures significantly associated with the outcome as potential risk factors. DeepEXPOKE-PRSKO and DeepEXPOKE-statKO models significantly outperform the other machine learning methods (p-value= 3.4e-01, 9.9e-05, and 8.1e-02 from two-sample t tests when DeepEXPOKE-PRSKO is compared with DeepEXPOKE-statKO, XGBoost, and Random Forest, respectively). In terms of selected variables, all four machine learning methods identify unique sets of exposures with unique findings being most prevalent (**Fig. 3B**). Interestingly, the findings from DeepEXPOKE-PRSKO and DeepEXPOKE-statKO diverged substantially, similar to how they differed from the other machine learning methods. Given that both models share the same underlying deep neural network (DNN) architecture, with the only variation being the use of knockoff variables, this suggests that the DNN is both sensitive enough for robust predictions and flexible enough to effectively address various challenges in exposome data analysis, such as genetic predispositions or collinearity. To further explore how the choice of knockoff affects model performance, we compared DeepEXPOKE-PRSKO and DeepEXPOKE-statKO across multiple performance metrics, including accuracy, sensitivity, and specificity. In all metrics, DeepEXPOKE-PRSKO consistently outperformed DeepEXPOKE-statKO (**Fig. 3C, D, E**), indicating that accounting for genetic predisposition is as crucial in identifying significant risk exposures for sepsis as accounting for exposure collinearity.

**Figure 3:**
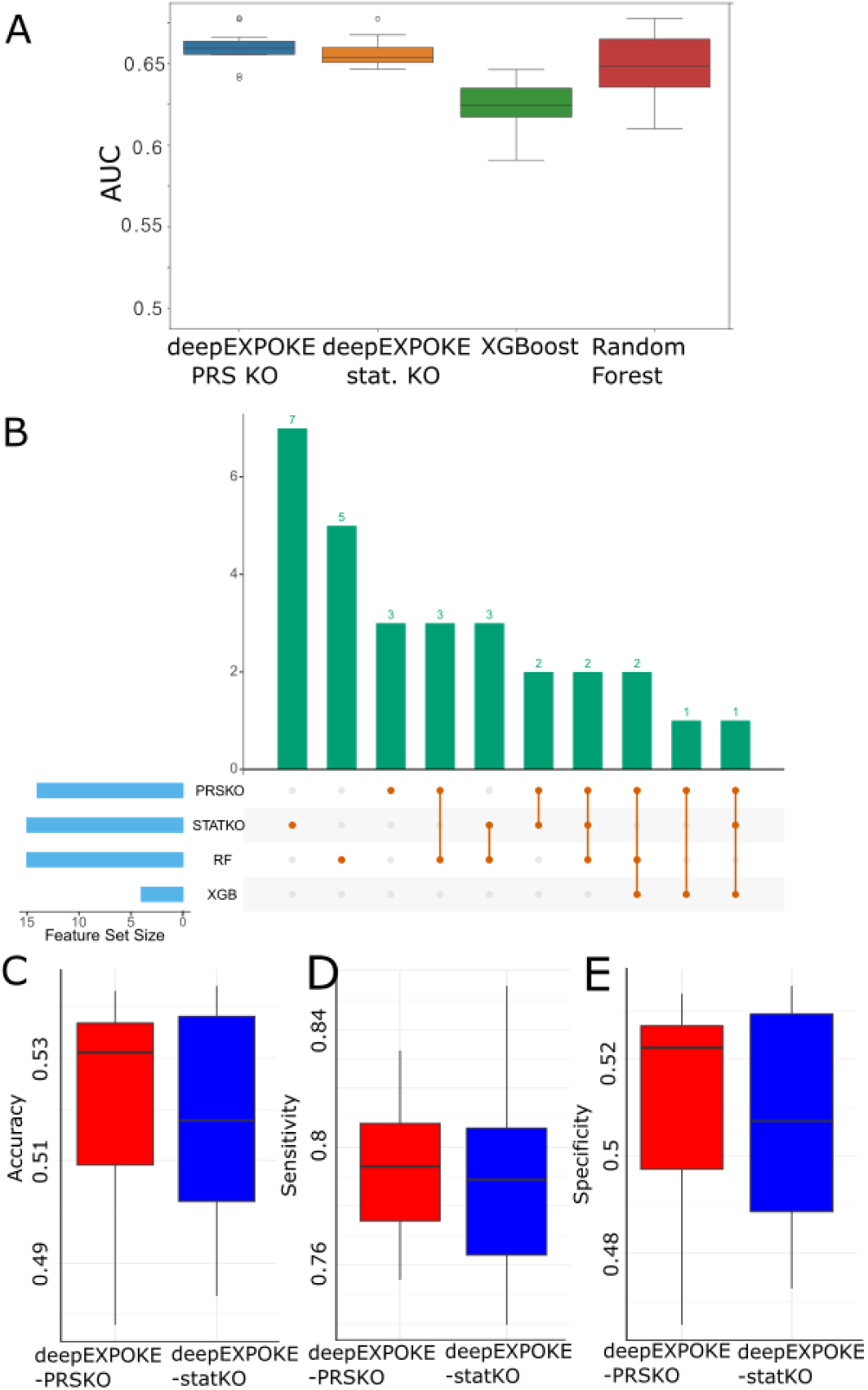
DeepEXPOKE accurately predicts outcomes by selecting important exposures. **A)** Boxplot showing the distribution and comparison of AUC values for models using DeepEXPOKE-PRS, DeepEXPOKE-STATKO, XGBoost, and Random Forest. Horizontal lines inside the bars denote median values. **B)** UpSet plot of identified selected features across DeepEXPOKE-PRS, DeepEXPOKE-STATKO, XGBoost, and Random Forest models. **C)** Boxplot comparison of accuracy between DeepEXPOKE-PRSKO (red) and DeepEXPOKE-STATKO (blue). **D)** Boxplot comparison of sensitivity between DeepEXPOKE-PRSKO (red) and DeepEXPOKE-STATKO (blue), **E)** Boxplot comparison of specificity between DeepEXPOKE-PRSKO (red) and DeepEXPOKE-STATKO (blue).

### DeepEXPOKE identifies risk factors that affect outcomes at different biological levels

To demonstrate how DeepEXPOKE-PRSKO and DeepEXPOKE-statKO differentiate risk exposures across biological layers, we estimated the effect size of the 14 and 15 exposures, respectively, that are strongly associated with sepsis incidence (**Fig. 4A, 4B**, see Methods). Both models identified five common exposures as risk factors (**Fig. 4C**), suggesting that these five shared exposures likely reflect the exposure risk independent of the genetic effect and collinearity (**Fig. 1C**). For example, glucose, triglycerides, and diabetes are known to exert the effect primarily at the metabolomic level^35–37^. Since metabolite levels and diabetes are highly responsive to environmental factors^38^, the results support our findings as independent of genetic effect and collinearity.

**Figure 4:**
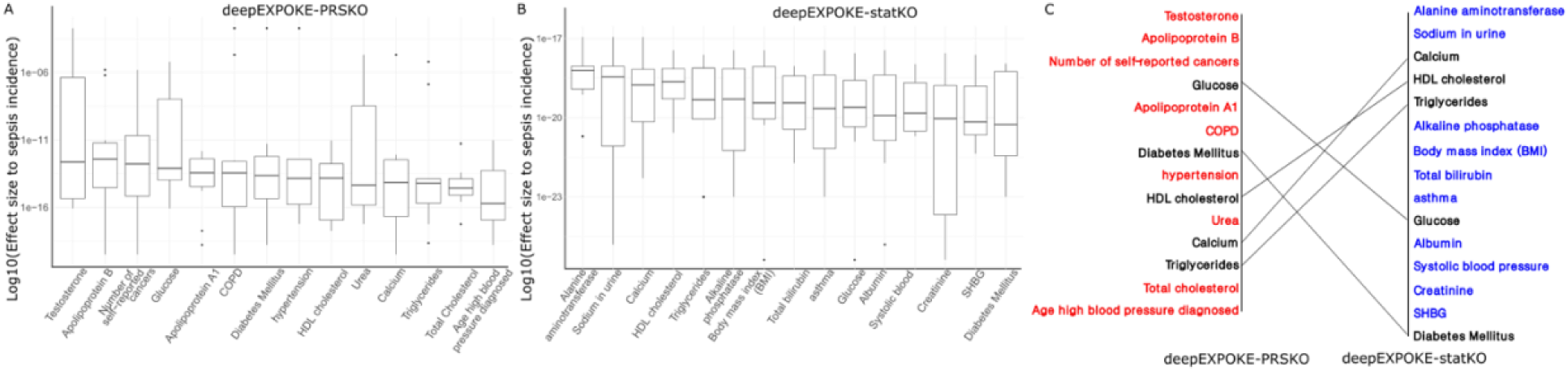
DeepEXPOKE identifies risk factors that affect outcomes at different biological levels. **A)** Boxplot showing the log-transformed effect sizes (W-statistics) of input risk factors on sepsis incidence using deepEXPOKE-PRSKO. Boxes are sorted by descending mean effect size value. **B)** Boxplot showing the log-transformed effect sizes (W-statistics) of input risk factors on sepsis incidence using deepEXPOKE-STATKO. Boxes are sorted by descending mean effect size value. **C)** Bump chart comparing significant risk factors between deepEXPOKE-PRSKO (red text) and deepEXPOKE-STATKO (blue text). Overlapping factors contributing to sepsis incidence in both models are noted in black text.

Separately, some exposures are identified only by DeepEXPOKE-statKO, such as asthma. Since risk factors only by DeepEXPOKE-statKO are likely driven mainly by genetic factors (**Fig. 1C**), this can partially explain the discrepancy where genome-wide association studies link asthma to increased sepsis risk^39^ but patient data often shows a negative association^40^, suggesting that asthma itself is not a direct sepsis risk factor, but the genetic variants associated with asthma may contribute to the risk. Similarly, the unique findings of DeepEXPOKE-PRSKO can be attributed to other factors than the genetic or exposome effects, such as collinearity. For example, blood pressure is found as a sepsis risk factor only by DeepEXPOKE- PRSKO. Despite many excellent antihypertensive drugs designed for controlling blood pressure, it remains difficult to achieve the desired target blood pressure; this phenomenon has been described as the hypertension paradox^31^. Our results suggest that this may be because hypertension is operated in association with other exposure associations, and it is not effective to control the internal biological system with the antihypertensive drugs unless the associations are controlled.

To assess generalizability, we estimated the effect sizes of 13 and 15 exposures significantly associated with CHD incidence by DeepEXPOKE-PRSKO and DeepEXPOKE-statKO, respectively (**S. Fig. 3A, B, C**) (see Methods). Consistently, exposures identified by both models, such as hypertension and total cholesterol, indicate that the risk is at the exposure level neither at the genetic level nor by collinearity. Hypertension and total cholesterol, are well-established topics for its genetic-independent contributions in cardiovascular research, as listed in a recent review paper^41^, support our finding. Overall, these findings demonstrate that DeepEXPOKE, using both statistical and PRS knockoffs, can effectively delineate the biological layers through which exposures influence disease phenotypes.

### DeepEXPOKE reveals potential mechanisms of sepsis

To demonstrate the clinical relevance of the DeepEXPOKE findings, we applied one-sample Mendelian Randomization using the 2-Stage Least Squares (2SLS) method^42^ to the sepsis incidence data using the PRS scores generated by pgsc_calc as instrumental variables for each exposure (**Fig. 5A, B**). Comparing these results with those from DeepEXPOKE-PRSKO and DeepEXPOKE-statKO highlights several potential sepsis mechanisms. First, DeepEXPOKE-PRSKO and DeepEXPOKE-statKO identified several exposures related to lipoprotein and lipid metabolism, such as hypertension, Apolipoprotein B, and BMI, consistently with 2SLS, further supporting their involvement in sepsis. Second, DeepEXPOKE-PRSKO and DeepEXPOKE-statKO uncovered additional significant exposures related to metabolism not captured as highly prioritized features by 2SLS, such as testosterone, total cholesterol, and Apolipoprotein A1^43, 44^, providing a more comprehensive view of how the metabolism pathways are related to sepsis. Third, DeepEXPOKE-PRSKO and DeepEXPOKE-statKO allow us to differentiate the exposure effects by the genetic, exposure, or collinearity-related influences. While both models emphasize the role of lipoprotein and lipid metabolism in sepsis risk, DeepEXPOKE-PRSKO uniquely identified Apolipoprotein B, among others. As DeepEXPOKE-PRSKO controls for genetic influences, this exposure likely impacts sepsis risk through collinearity with other exposures. Indeed, recent genetic studies suggest that lower low-density lipoprotein (LDL) levels, primarily Apolipoprotein B, tend to be more associative than causal for increased mortality risk in sepsis^45^, further supporting and validating our findings. Fourth, by distinguishing between genetic and exposure influences, DeepEXPOKE can inform clinical trial designs for interventions targeting lipoprotein cholesterol, such as cholesteryl ester transfer protein (CETP) inhibitors or testosterone replacement therapy (TRT)^46^ for sepsis management. For instance, if Apolipoprotein B is only associatively linked to sepsis risk, patient selection for treatment could focus on other lipid metabolism features with strong effect sizes identified by both DeepEXPOKE-PRSKO and DeepEXPOKE-statKO, such as urea, testosterone, total cholesterol, and Apolipoprotein A1.

**Figure 5.**
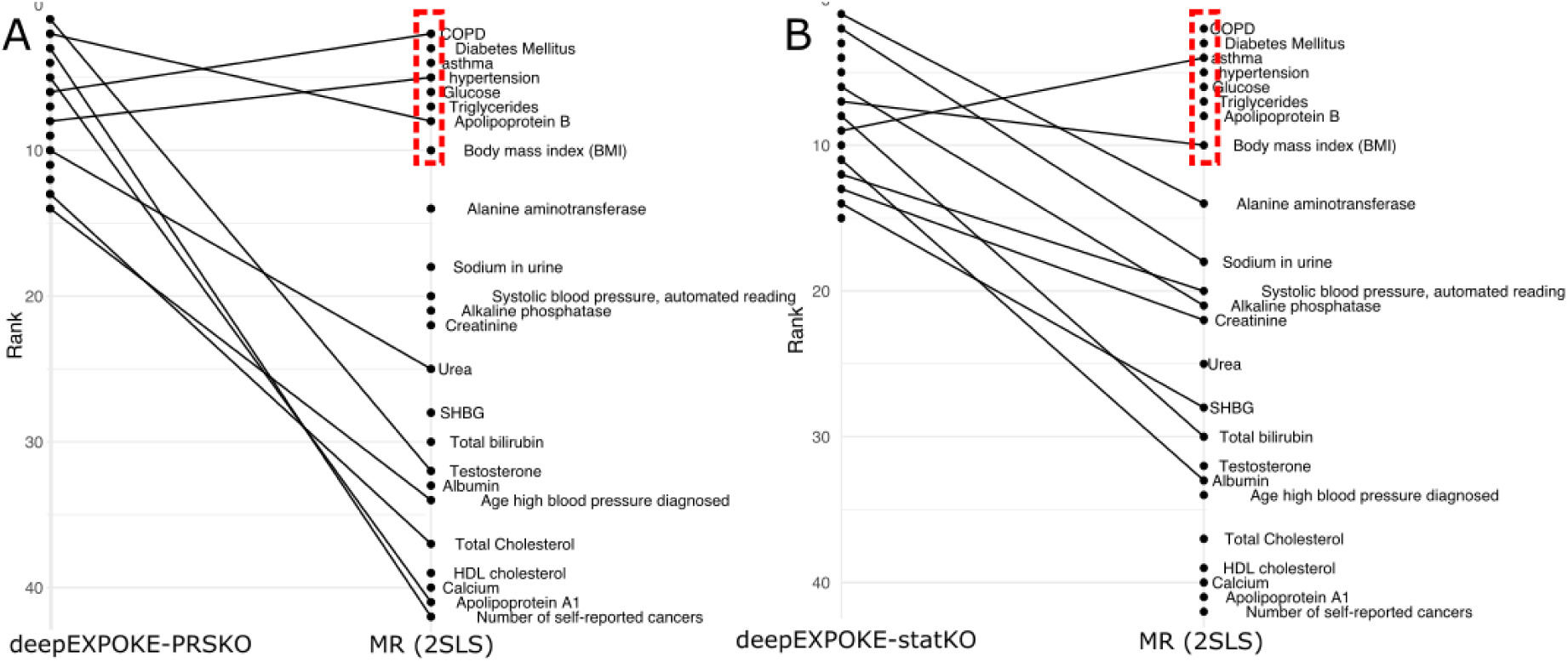
DeepEXPOKE reveals potential mechanisms of sepsis. **A)** Bump chart of selected risk factors between deepEXPOKE-PRSKO and MR with 2SLS. The red dashed box represents the eight highest ranking risk factors identified via MR. **B)** Bump chart of selected risk factors between deepEXPOKE- STATKO and MR with 2SLS. The red dashed box represents the eight highest ranking risk factors identified via MR.

## DISCUSSION

In this study, we developed DeepEXPOKE, a novel deep neural network (DNN) framework, to identify exposure risk factors for complex diseases by both employing statistical knockoff features and integrating polygenic risk score (PRS) as a novel type of biologically-driven knockoff. Our approach leverages the mathematical properties of PRS to validate it as a valid knockoff, thereby maintaining control over the genetic effects in exposome data analysis. Using these knockoffs, DeepEXPOKE successfully predicts disease outcomes by selecting and prioritizing important exposure variables that contribute to sepsis and coronary heart disease (CHD).

Further, DeepEXPOKE allows us to distinguish the genetic and exposome-level effects of the exposures, enabling a mechanistic understanding of sepsis and CHD. For sepsis, the exposome-level impacts of key metabolites, such as lipoproteins and glucose, were highlighted by the fact that both DeepEXPOKE- PRSKO and DeepEXPOKE-statKO identified them as risk factors. Also, deepEXPOKE helps explain the inconsistency of asthma genomics and patient data studies regarding its sepsis risk by estimating its risk only with statKO, but not with PRSKO. Since the risk estimated only by DeepEXPOKE-statKO indicates risk strictly from genetic factors, not from exposure collinearity nor exposure effect, the inconsistency confirms the identification of asthma only by our DeepEXPOKE-statKO method highlights how the separation of genetic and non-genetic effects helps elucidate the ongoing paradox in understanding the totality of effects^47, 48^, such as those related to asthma in our analysis. Similarly, for CHD, DeepEXPOKE- statKO and DeepEXPOKE-PRSKO distinctively identify a strong non-genetic effect of both hypertension and total cholesterol, and a strong genetic effect related to testosterone, specifying the biological layer at which these effects occur. Altogether, DeepEXPOKE clearly demonstrates that genetic risk alone does not fully capture an individual’s susceptibility to sepsis and CHD, aligning with findings that emphasize the multifactorial nature of sepsis and cardiovascular events^49^. This underscores the need for prevention and management strategies that account for both genetic and non-genetic risk factors.

DeepEXPOKE is also advantageous in its use of deep neural networks to capture nonlinear exposure effects. For example, DeepEXPOKE-PRSKO identifies testosterone as having a significant effect on sepsis risk while 2SLS, a linear model-based MR analysis, fails to highlight its importance. Testosterone is known to support immune functions, such as enhancing neutrophil differentiation and certain inflammatory responses, while also promoting an immunosuppressive phenotype that can inhibit bactericidal properties^28^. This contradictory behavior suggests a non-linear relationship where varying levels of testosterone may differentially influence immune cell behavior in sepsis.

Despite its advantages in risk factor identification and prediction capabilities, DeepEXPOKE is limited in a few aspects. First, while PRS can be useful, some data complexities exist in the use of PRS as instrumental variables, such as pleiotropy, population stratifications, and linkage disequilibrium (LD).

Pleiotropy occurs when a single genetic variant influences multiple traits through different biological pathways. In the context of PRS, this means that the score may be associated with the outcome through pathways involving other than the exposure of interest, thus violating the exclusion restriction assumption of instrumental variables. Population stratification, which involves differences in allele frequencies and trait distributions between subpopulations due to ancestry, can confound PRS-outcome associations if not properly controlled. Additionally, LD, the non-random association of alleles at different loci, can cause the PRS to overestimate the genetic component of the exposure when constructed from closely linked variants. Although we employed the PRS method developed to address ancestry structure and LD^32^, unmeasured portion of the data complexities may still persist. Second, the model assumes that the causal direction flows from exposure to outcome, whether through genetic, non-genetic, or collinearity effects. However, in the case of collinearity, this assumption may not hold true. It is important to distinguish correlation due to causation from other forms of association, such as reverse causation or confounding. Reverse causality occurs when an outcome influences the exposure, rather than the exposure influencing the outcome. For instance, if disease severity (outcome) affects lifestyle behaviors (exposure), you would observe a correlation between lifestyle and disease, which might misleadingly suggest that lifestyle changes lead to changes in disease severity. Finally, bidirectional relationships can complicate causal relationship interpretation. For example, obesity and depression may influence each other: obesity can lead to depression, and depression can lead to obesity. This reciprocal relationship can be detected through the presence of correlation, but it complicates the interpretation of causality^50^. Thus, careful interpretation is warranted to understand the collinear effect of exposures.

The findings of this study have significant implications for understanding and managing complex diseases such as sepsis and CHD. By developing DeepEXPOKE, a deep neural network framework incorporating both the well-established statistical knockoff and a novel polygenic risk score (PRS) knockoff, we have demonstrated a powerful tool for identifying critical exposure risk factors that contribute to disease outcomes at the genetic, non-genetic, or collinearity level.

## Methods

### Calculating polygenic risk scores

PRS for each of our input exposures in our UK Biobank patient data were calculated using pgsc_calc software. Pgsc_calc enables streamlined calculation of PRS using a structured workflow system called Nextflow. The tool is a bioinformatics best-practice analysis pipeline for calculating polygenic risk scores on samples with imputed genotypes using existing scoring files from the Polygenic Score (PGS) Catalog. In the first step of the calculations, we used existing score files with comprehensive variant information from two UK Biobank PRS evaluation studies^51, 52^, and then matched these score files to target genomes downloaded from the UK Biobank in VCF format. Genomic data and score files are in the GRCh37 specified genome build. Variants in the scoring files were matched against variants in the target genome data. Finally, we performed a scoring step that calculated PRS for each of our samples across our 42 features as a linear sum of weights and dosages. Pgsc_calc software leverages utilities from pgscatalog_utils and PLINK2 as part of its scoring pipeline, enabling us to compare PRS calculation differences between conventional methods (PLINK2) and the complete pgsc_calc pipeline.

### Exposures for sepsis and CHD in the UKBB

We identified sepsis cases in the UK Biobank using The International Classification of Diseases, Tenth Revision (ICD-10) codes. The codes we included were A02.1, A22.7, A26.7, A32.7, A40, A40.0, A41.1, A41.2, A41.3, A41.4, A41.5, A41.6, A41.9, A42.7, B37.7, and O85, corresponding to septicemia subtypes ‘Salmonella Septicemia,’ ‘Anthrax Septicemia,’ ‘Erysipelothrix Septicemia,’ ‘Listerial Septicemia,’ ‘Streptococcal Septicemia,’ ‘Other Septicemia,’ ‘Unspecified Septicemia,’ ‘Actinomycotic Septicemia,’ ‘Candidal Septicemia,’ and ‘Puerperal Sepsis.’ Sepsis deaths were confirmed using the corresponding ICD- 10 codes from death cause records. For our control subjects, we selected a sample size of 79,791 individuals with no recorded hospital diagnoses of any sepsis or sepsis subtype.

42 unique features in the UK Biobank were selected as input variables for our PRS scoring framework and subsequent causal analyses. These variables fit into the following categories of measurements: routine measures of cardiac activity, respiratory condition, blood and urine biomarkers, anthropometry, lifestyle and environment, family history, complex disease, and health and medical history. All complex diseases were categorically coded with 0 representing “no disease” and 1 representing “diseased” states.

### DeepEXPOKE Causal Association Estimation

To identify causal associations between the selected 42 features and the derived phenotypes, we built the DAG-deepVASE DNN architecture consisting of an input layer with twice the number of neurons as input features, accommodating both the original features and knockoff features in a pairwise manner. The network has two hidden layers, each with the same number of neurons as the input variables and uses the rectified linear unit (ReLU) activation function. The weights in the network are initialized using the Glorot normal initializer, and L1 (LASSO) regularization is applied to prevent overfitting. The model is optimized using the Adam algorithm with a learning rate of 0.001, and the mean squared error (MSE) serves as the loss function during training^23^.

For identifying linear associations, we implemented Lee and Hastie’s log-likelihood model to evaluate all possible pairs of variables. Lee and Hastie’s log-likelihood model provides a framework for learning the structure of mixed graphical models that include both continuous and discrete variables^53^. It evaluates conditional dependencies between variables using Gaussian regression for continuous variables and multiclass logistic regression for discrete variables. By applying a log-likelihood approach with a sparsity penalty, this model allows for the identification of important variable associations and, by eliminating non-significant edges, results in a simpler interpretable model^23^. The sparsity penalty parameter was set to 0.3, allowing us to select the most important variables. The variable pairs that remained significant after applying the sparsity penalty were identified as linear associations.

To identify nonlinear associations between input variables *x*_*i*_, DAG-deepVASE constructs a series of perceptron layers between *X*_∖*i*_ = {*x*_1_, *x*_2_, …, *x*_*i*−1_, *x*_*i*+1_, …, *x*_*M*_} and *x*_*i*_. This allows us to estimate the effect size of the association between *x*_*j*_ ∈ *X*_∖*i*_ and *x*_*i*_ using model-X knockoffs. For the input variables *x*_*i*_ and *x*_*j*_, the method ensures exchangeability 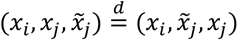, where 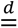 denotes equality in distribution. This property allows the identification of causal relationships, differentiating them from simple observed correlations. For instance, if (*x*_*i*_, *x*_*k*_) is a correlation without a causal link, then the feature exchangeability 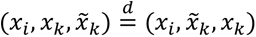 will hold, making the relationship measures |*R* _*ik*_ | and |*R̃*_*ik*_ | symmetric and exchangeable around zero where *R* _*ik*_ is the correlation between the original feature *X*_*i*_ and the outcome after training, and *R̃*_*ik*_ is the correlation between the knockoff *X̃*_*i*_ and the outcome after training. On the other hand, if (*x*_*i*_, *x*_*j*_) is a causal relationship, *S* _*ij*_ = |*R* _*ik*_| − |*R̃*_*ik*_| will capture the deviation of this relationship from the null hypothesis. The knockoff matrix *X̃* is designed to match the correlation structure within *X* while minimizing cross-correlation with the outcome variable *Y* ^23^.

The knockoff variables *X̃* = (*x̃*_1_, …, *x̃*_*p*_)^*T*^ follow two main properties: exchangeability, where 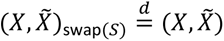, where swapping denotes exchanging any subset *S* ⊆ {1, …, *M*} of the variables *x*_*j*_, and independence, meaning *X̃* ⊥ *Y* |*X*, ensuring that *X̃* is independent of *X* given the outcome *Y* .

To construct knockoffs, we assume *X* ∼ *N*(0, Σ), with *Σ* ∈ ℝ^*M*×*M*^as the covariance matrix. A valid knockoff construction for *X̃* is given by:

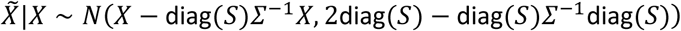

The effect size *S* _*ik*_ is computed as the difference in correlation strength between the original feature and the knockoff feature:

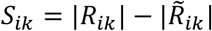

If the original feature has a stronger correlation than its knockoff counterpart, it is called as a selected feature for continued analysis. Effect size calculations for each identified association involved pairing each input variable with its knockoff counterpart and using matrix operations across the DNN’s layers to quantify the importance of each original variable relative to its knockoff. The effect size determining process utilizes weight matrices to compute feature importance scores. These importance scores are then used to calculate an effect size for each feature that reflects the strength of each feature’s association with the outcome of interest, calculated as the difference between the importance of the original feature and its knockoff variable^23^. We parameterized *q* as a user-defined nominal false discovery rate (FDR), and subsequently set FDR to a level *q* = 0.05 to enable balance estimation of associations.

### Comparison of DeepEXPOKE to Other Machine Learning Methods

To evaluate DeepEXPOKE-PRS’s performance in a comparative manner, we first tested it against DeepEXPOKE-statKO and compared accuracy, sensitivity, and specificity metrics between the two. A two-sample t-test was performed to compare the difference in mean values of each of the three metrics.

Next, we compared model area under the receiver operating curve (AUC) values for DeepEXPOKE-PRS DeepEXPOKE-statKO, XGBoost, and Random Forest. XGBoost builds a series of decision trees sequentially, where each new tree recursively corrects errors made by previous trees ^33^. Random Forest, rather than sequentially building trees, builds several independent decision trees and then aggregates their outputs to yield one resultant outcome^54^. XGBoost, Random Forest, and KNN parameter assignment and model setup were conducted using Python 3.8.5 and the *scikit-learn* and *xgboost* software packages^55^.

For comparing the overlap of selected features across different machine learning methods, we used the *UpSetR* package to visualize the intersecting sets of features between DeepEXPOKE-PRS, DeepEXPOKE- Stat, XGBoost, Random Forest, and linear regression machine learning methods ^56^. XGBoost’s feature selection was performed automatically within the framework of the algorithm. Random Forest feature selection was done via a permutation feature importance threshold model. The mean value was the threshold for which selected features were identified. After the model was fit, features that had non-zero coefficients were extracted and identified as selected features.

### One-Sample Mendelian Randomization (MR) Analysis

For the primary Mendelian Randomization (MR) analysis, we used the calculated PRS generated from the pgsc_calc software as instrumental variables (IVs) to estimate relationships between our set of 42 exposures and the derived phenotypes of interest^32^. We first mathematically validated that PRS were appropriate instrumental variables to use as part of the MR step. The PRS generated by pgsc_calc for each feature serve as IVs for each exposure in our analysis, and PRS were used to capture the genetic predisposition to the exposures in a similar manner to the way single nucleotide polymorphisms (SNPs) are conventionally used in MR analysis^13^.

We used the two-stage least squares (2SLS) method for MR, a standard approach for one-sample MR experiments. The 2SLS approach involves two primary steps. First, the exposure (one of the 42 features) is regressed on the IV (feature PRS). This stage aims to estimate the genetically driven predicted values of the exposure. Second, the outcome is then regressed on the predicted values of the exposure calculated from the first stage. The causal effect of the exposure on the outcome is given by the coefficient of the genetically predicted exposure from the stage-two regression. We implemented the 2SLS approach using *ivreg* from the AER package in R. For each exposure, we fit separate models, using the exposure’s corresponding PRS as the IV. Robust standard errors were used to account for any heteroscedasticity. All statistical analyses were conducted using R version 4.2.1.

## Data Availability

All data produced in the present study are from the UK Biobank. Polygenic risk score scoring files can be found online at: https://www.pgscatalog.org/

---

**S. Figure 1.**
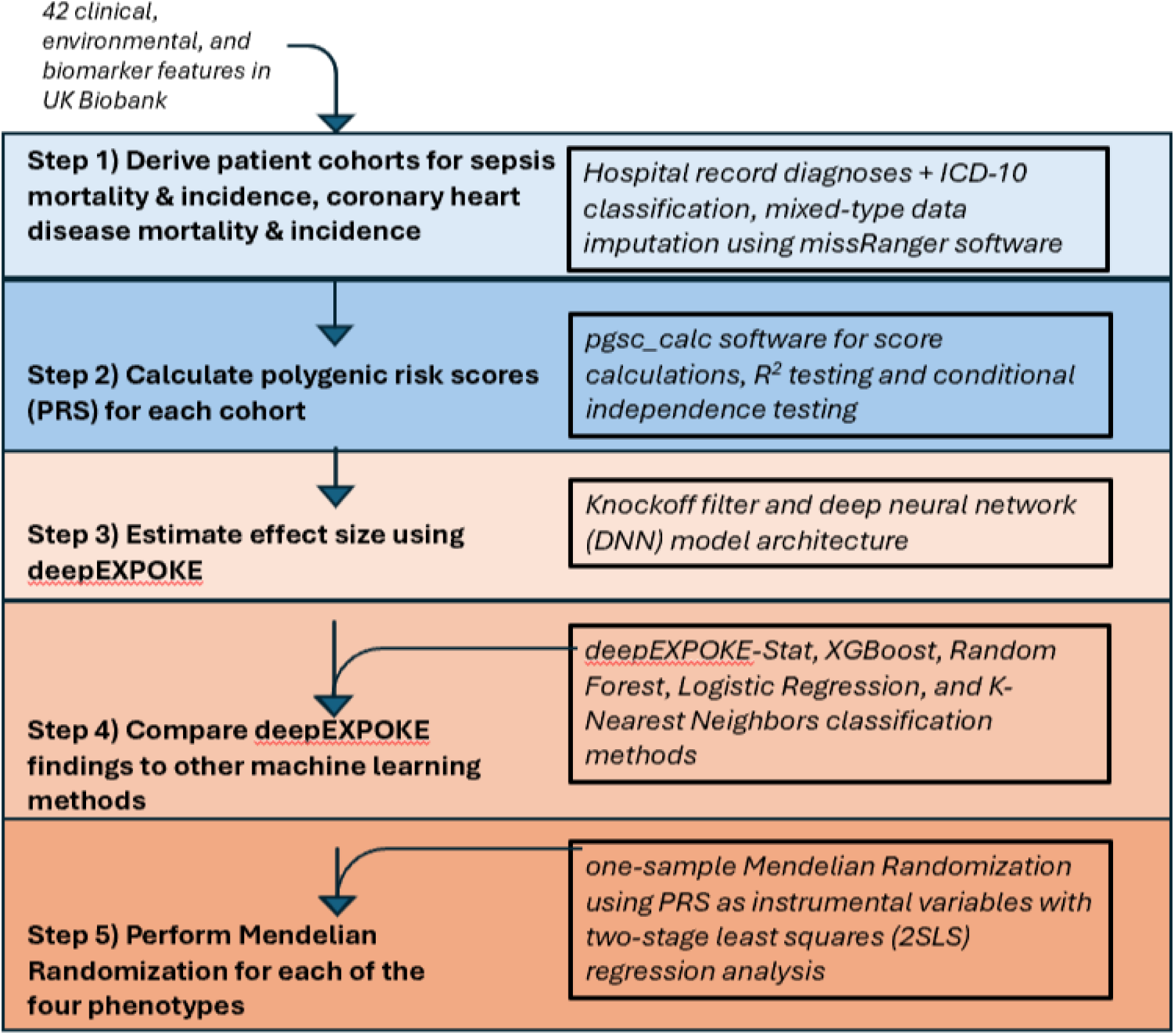
Overview of experimental design: cohort establishment, polygenic risk score calculations, DeepEXPOKE model setup, performance evaluation and comparison to other machine learning methods, and Mendelian Randomization setup.

**S. Figure 2.**
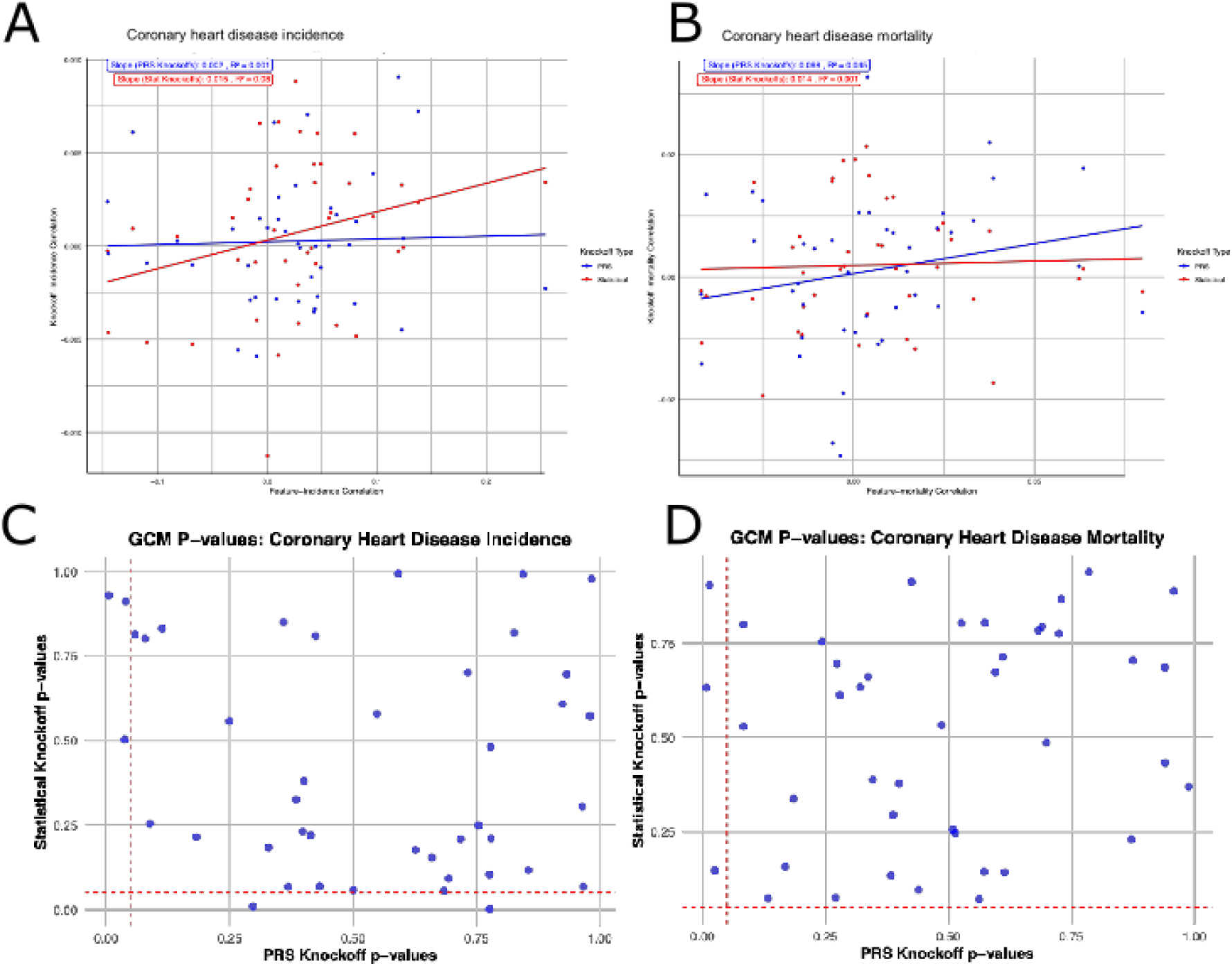
**A)** Scatterplot of correlation values for knockoff variables and CHD incidence plotted against the correlation values for input exposures and CHD incidence. **B)** Scatterplot of correlation values for knockoff variables and CHD mortality plotted against the correlation values for input exposures and CHD mortality. In A) and B), the blue points and regression line represent the data for PRS knockoffs, and the red points and regression line represent the data for statistical knockoffs. **C)** P-values for the GCM test for conditional independence for the sepsis incidence phenotype for PRS knockoffs (x-axis) and statistical knockoffs (y-axis). **D)** P-values for the GCM test for conditional independence for the sepsis mortality phenotype for PRS knockoffs (x-axis) and statistical knockoffs (y-axis). In C) and D), red dashed lines represent significance levels of 0.05. Points above the horizontal red dashed line exhibit conditional independence in the construct of PRS knockoffs, and points to the right of the vertical red dashed line exhibit conditional independence in the construct of statistical knockoffs.

**S. Figure 3.**
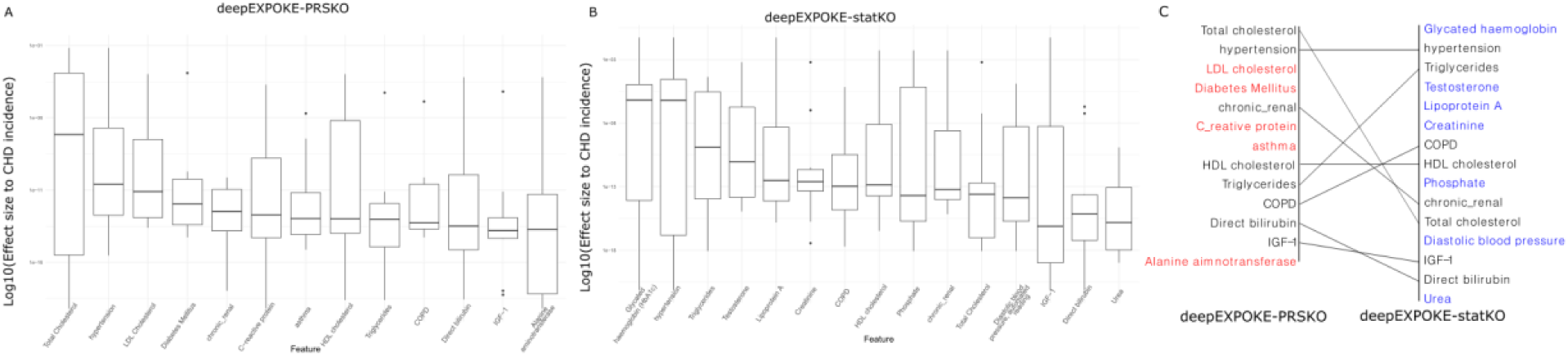
**A)** Boxplot showing the log-transformed effect sizes (W-statistics) of input risk factors on sepsis incidence using deepEXPOKE-PRSKO. Boxes are sorted by descending mean effect size value. **B)** Boxplot showing the log-transformed effect sizes (W-statistics) of input risk factors on sepsis incidence using deepEXPOKE-STATKO. Boxes are sorted by descending mean effect size value. **C)** Bump chart comparing significant risk factors between deepEXPOKE-PRSKO (red text) and deepEXPOKE-STATKO (blue text). Overlapping factors contributing to sepsis incidence in both models are noted in black text.

